# Retraining Gastrocnemius Muscle Coordination Reduces Late-Stance Knee Contact Force in Individuals with Knee Osteoarthritis

**DOI:** 10.64898/2026.01.14.26344117

**Authors:** Michelle R. Joyce, Julie Muccini, Benjamin Randoing, Scott L. Delp, Scott D. Uhlrich

## Abstract

Musculoskeletal simulations and experiments in young adults without knee pain have demonstrated that reducing gastrocnemius muscle activity can reduce knee contact force, which may reduce pain and progression of knee osteoarthritis. This study investigated whether individuals with knee osteoarthritis could reduce gastrocnemius electromyography (EMG) when provided with haptic biofeedback and consequently reduce late-stance knee contact force. Individuals with tibiofemoral osteoarthritis walked on a treadmill with adaptive biofeedback instructing them to reduce gastrocnemius EMG. Thirteen of eighteen participants reduced their average gastrocnemius EMG by at least 10% during an initial 30-minute training session and thereby qualified for a second session. During the second session, participants received the same biofeedback, and we estimated knee contact force using musculoskeletal models and static optimization. With feedback, participants reduced gastrocnemius EMG by 25±15% (p<0.001) and reduced the late-stance peak of knee contact force by 12%, or 0.38±0.47 times body weight (p=0.01). However, the average EMG of the vasti muscles increased by 38±34% (p=0.004), which contributed to an increase in early-stance knee contact force in some participants. Ten out of thirteen reduced their knee contact force impulse with the feedback. While additional work is needed to mitigate increases in vasti EMG, our data demonstrates that individuals with knee osteoarthritis can reduce gastrocnemius EMG and late-stance knee contact force in a brief period of training, suggesting the potential of muscle coordination retraining as a non-surgical intervention for knee osteoarthritis.

## I. Introduction

IN individuals with knee osteoarthritis, high knee joint loads are associated with cartilage degeneration [1], [2]; thus, reducing joint loading during daily activities, such as walking, is an interventional target. During walking, the knee joint experiences forces two to four times body weight (BW), and 50–75% of knee contact force is generated by the muscles that span the knee [3], [4], [5]

Gait retraining studies have investigated methods of adjusting kinematics, such as the foot progression angle [6], [7], [8], [9] and trunk sway [10], [11], to reduce the knee adduction moment, a measure that represents the mediolateral distribution of the intersegmental knee contact force. However, reductions in the knee adduction moment do not always result in reductions in medial knee contact force [12], [13]. Additionally, since changing the knee adduction moment redistributes forces between the medial and lateral compartments, individuals with osteoarthritis in both compartments are not candidates for interventions that target the knee adduction moment. Directly targeting the muscle force contribution to knee contact force provides an alternate way to reduce forces in both tibiofemoral compartments.

The contributions of different muscles to knee contact force varies across the stance phase of walking [3], [14]. The quadriceps contribute most to muscle-generated knee contact force in early-stance, and the gastrocnemius dominates in late-stance [5]. Musculoskeletal modeling and simulations can identify strategies to reduce the muscle contributions to knee contact force while walking. These simulations have suggested that reducing gastrocnemius muscle activation during walking can dramatically reduce the late-stance peak of knee contact force without requiring changes in gait kinematics [15], [16], [17].

Uhlrich et al. demonstrated that young adults without knee osteoarthritis can reduce their gastrocnemius activity and knee contact force while walking with visual feedback about their muscle activity [14]. However, it is unknown whether individuals with knee osteoarthritis would also be able to learn this coordination change and achieve similar reductions in knee loading, due to age, pain, or changes in gait or muscle coordination [18], [19], [20]. Haptic feedback can be integrated into a wearable device, clothing, or footwear, making it well suited for gait retraining in natural environments [21]. Yet it is unknown whether haptic feedback can be used to retrain muscle coordination patterns while walking.

The objective of this study was to investigate whether individuals with knee osteoarthritis can reduce late-stance knee contact force using haptic feedback about the average electromyography (EMG) of their gastrocnemius. We hypothesized that individuals with knee osteoarthritis can reduce their gastrocnemius EMG while walking with real-time haptic feedback about their gastrocnemius activity. Furthermore, we hypothesized that individuals who were able to reduce their gastrocnemius EMG would reduce their late-stance knee contact force peak. As an exploratory aim, we investigated whether the feedback elicited changes in sagittal plane kinematics, kinetics, and the average EMG of other knee-spanning muscles.

## II. Methods

### A. Experiment

Eighteen adults with tibiofemoral knee osteoarthritis (Table 1) participated in this study after providing informed consent to a protocol approved by Stanford University’s Institutional Review Board (IRB-34928). This study was registered on ClinicalTrials.gov (NCT06208631). Individuals were included if they had physician-diagnosed knee osteoarthritis in the medial and/or lateral compartments, could walk for at least 60 minutes without aid, and reported a typical pain level of ≤4 on a 0–10 numeric rating scale (i.e., only mild pain). Exclusion criteria included arthritis in other lower limb joints, a history of rheumatoid arthritis, previous joint replacement, and body mass index (BMI) greater than 35.

**TABLE I.** Participant Characteristics.

|  | Session 1<br>(n=18) | Session 2<br>(n=13) |
| --- | --- | --- |
| Age (years) | 59.4 [14.6] | 57.6 [17.0] |
| Height (m) | 1.72 [0.12] | 1.75 [0.12] |
| Weight (kg) | 75.8 [15.6] | 78.2 [17.4] |
| Walking Speed (m/s) | 0.99 [0.18] | 0.99 [0.19] |
| Sex | 12F / 6M | 7F / 6M |
| Kellgren-Lawrence<br>Grade |  | I: 1; II: 6;<br>III: 4; IV: 1 |

Values are shown as mean [standard deviation]
Kellgren-Lawrence Grade is not reported for one participant.

#### 1) Session One

Participants completed one or two laboratory sessions, depending on their ability to modify the coordination of their muscles during the first session. The first session aimed to determine if each participant could reduce their gastrocnemius EMG by at least 10% relative to their natural walking. The 10% threshold was expected to be attainable with a gait modification, avoid noise in the EMG signal, and affect the late-stance peak of knee contact force. During this session, participants acclimated to walking on the treadmill, received verbal suggestions to aid in reducing gastrocnemius EMG, and began the coordination retraining during three to five six-minute trials with haptic biofeedback.

We placed wireless surface EMG sensors (Trigno Avanti Sensors, Delsys Inc., Natick, MA, USA) on the medial gastrocnemius and soleus of the leg with osteoarthritis, or in individuals with bilateral knee osteoarthritis, on the leg with greater knee pain. The EMG signal for each muscle was processed by bandpass filtering (30-500 Hz, 4th order, zero-phase shift Butterworth), rectifying, and low-pass filtering (6 Hz, 4th order, zero-phase shift Butterworth). EMG signals during the walking trials were normalized by the maximum value measured for each muscle during standing calf raises and resisted seated calf raises.

Participants then walked at a self-selected speed on a splitbelt force-instrumented treadmill (Bertec Corporation, Columbus, OH, USA) until they felt comfortable walking on the treadmill (minimum 8 minutes). The treadmill speed was set to a pace that participants felt they could consistently maintain for one hour. Treadmill speed remained constant for all subsequent trials and sessions. We recorded two additional minutes of natural walking as a baseline trial, and the average medial gastrocnemius EMG during the stance phase of the trial was used as the baseline value for this session. Next, we provided strategy suggestions to aid with the initial training (described in the Supplementary Material). Participants were not constrained to follow these suggestions and were encouraged to explore various walking strategies to identify the strategy that most effectively achieved the feedback objective while allowing them to walk as naturally as possible.

During the feedback trials, individuals received unilateral haptic biofeedback after each step, instructing them to walk with less gastrocnemius activity. Force plate and EMG data were streamed from the motion capture software (Cortex v9.0, Motion Analysis Corporation, Santa Rosa, CA, USA) to custom MATLAB software (MathWorks Inc., Natick MA, USA) for real-time analysis and feedback. For each step, the filtered, normalized gastrocnemius EMG signal was averaged over stance phase and compared to the baseline value. If the step’s average EMG was greater than 95% of the baseline value, the participant received two vibrations (Fig. 1). If the step’s EMG decreased from baseline between 5% and the trial’s adaptive goal, participants received one vibration. Receiving no vibrations indicated that the step’s EMG decreased below the goal. Therefore, the overall objective was to make gait adjustments that consistently eliminated the vibrations. The adaptive feedback goal started at a 10% decrease from baseline. If the average change over a 50-step window during the trial exceeded the goal, it was lowered by 10% for the next trial, until a maximum goal of 30% reduction from baseline. Although feedback was given on one leg, participants were encouraged to walk symmetrically.

**Fig. 1.**
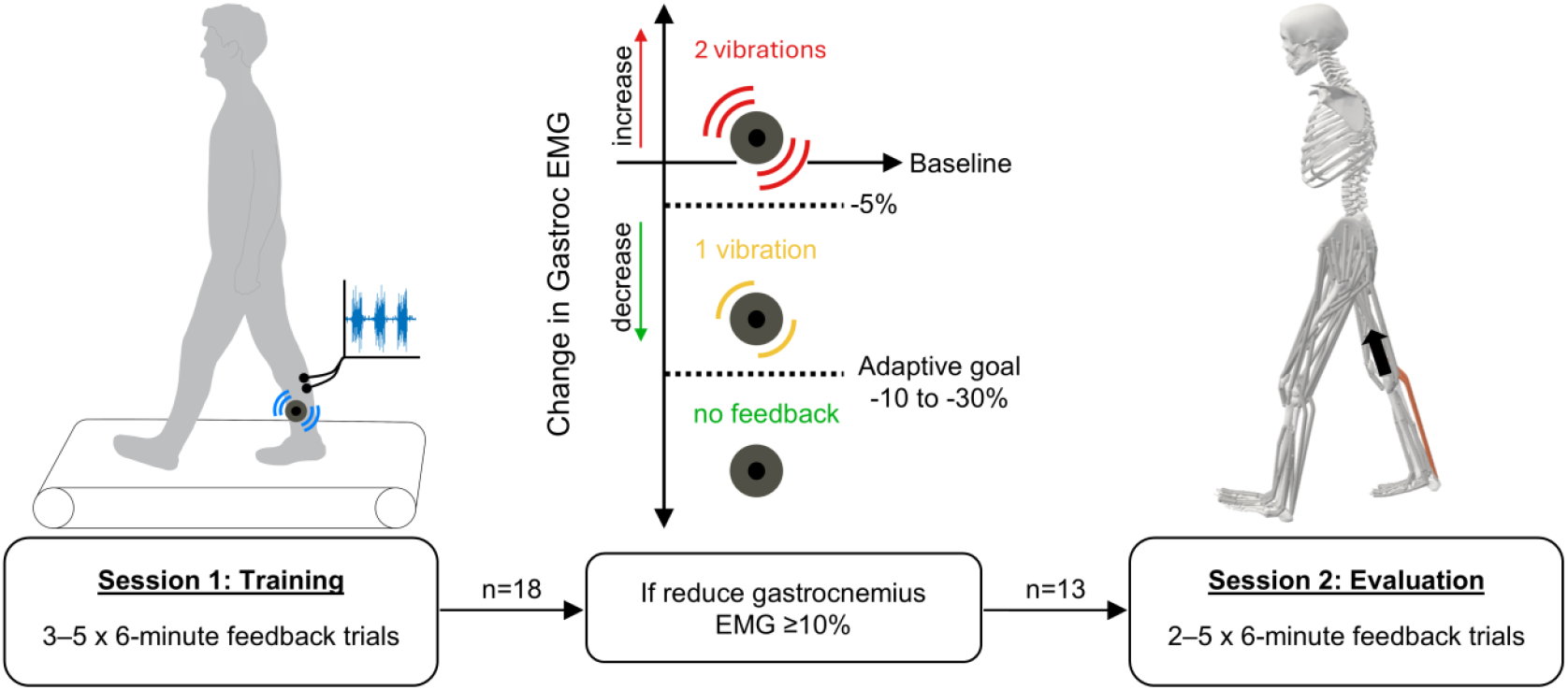
Study Overview. During the first session, 18 adults with knee osteoarthritis walked on a force-instrumented treadmill and began training to reduce their gastrocnemius activity, which was monitored through surface electromyography (EMG). Participants received vibrational haptic feedback after each step, with the number of vibrations corresponding to the change in gastrocnemius EMG from baseline. The objective was to receive no feedback by reducing gastrocnemius EMG below an adaptive goal. If participants achieved an average reduction of 10% from baseline over 50 consecutive steps within 5 feedback trials (30 minutes of training), they qualified for the second training session. Thirteen participants qualified and continued training to reduce their gastrocnemius EMG with the same feedback. Data from this session informed musculoskeletal models to estimate knee contact force.

During the first session, participants completed between three and five six-minute feedback trials. If they were able to achieve an average 10% reduction in gastrocnemius activity over 50 consecutive steps, they could finish the session after three trials. If not, participants would continue until they achieved the 10% average reduction or for a maximum of five trials. Participants who achieved the 10% reduction returned two–seven days later for a second session. Those who did not achieve the 10% reduction after five trials did not continue to the second session.

#### 2) Session Two

Thirteen participants qualified for the second gait retraining session (Table 1). We obtained x-rays for twelve participants and Kellgren-Lawrence (KL) grades were determined by KneeNet, an automated tool to assess knee osteoarthritis severity [22].We could not obtain an x-ray and severity grade for one individual, who had a physician diagnosis of mild osteoarthritis based on symptoms and magnetic resonance imaging (MRI). During the second session, we collected motion capture data and surface EMG data from muscles on both legs. Reflective markers were placed on the second and fifth metatarsal heads, calcanei, medial and lateral malleoli, medial and lateral femoral epicondyles, anterior and posterior superior iliac spines, acromion processes, sternum, and C7 vertebrae. Additional marker clusters were placed on the thighs and shanks for segment tracking. We collected a static pose for model scaling and a dynamic hip range-of-motion trial for calculating hip-joint centers [23]. We placed EMG electrodes bilaterally on the medial gastrocnemius, soleus, vastus medialis, and biceps femoris and unilaterally on the feedback leg’s lateral gastrocnemius, tibialis anterior, vastus lateralis, rectus femoris, and semitendinosus. After a warm-up of at least three minutes of treadmill walking, participants performed a series of maximum voluntary contraction activities. These included resisted isometric knee flexion, ankle dorsiflexion, hip flexion, body-weight squats, standing calf raises, resisted seated calf raises and isokinetic knee extension. EMG signals during the walking trials were normalized by the maximum value measured for each muscle during these maximum voluntary contraction activities.

Participants walked at their constant self-selected speed for three minutes to reacclimate to walking on the treadmill, followed by a new two-minute baseline trial. Prior to the feedback trials, we reviewed the feedback instructions and strategy suggestions. During this session, participants completed between two and five six-minute feedback trials. The target to finish the session early was lowered to a 20% average decrease from baseline over 50 consecutive steps. Participants began with the adaptive feedback goal set at 10%, or 20% if they advanced to a 30% goal in the first session. The goal was lowered if a 50-step average throughout the trial exceeded the goal. Participants continued feedback trials until they achieved the 20% threshold or completed a maximum of five trials. Data collected in this session was used to inform a musculoskeletal modeling workflow to calculate knee contact force.

### B. Data Analysis

To estimate joint kinematics, moments, and contact forces, we simulated the experimental data from the second session in OpenSim 4.5 [24], [25]. We used the musculoskeletal model from Rajagopal et al. [26], with modifications to passive muscle forces and hip abductor muscle paths [14]. First, we scaled the model using the static trial to match the participants’ anthropometry. We used OpenSim’s Inverse Kinematics tool to solve for joint kinematics. We then filtered the kinematics (6 Hz, 6th order, zero-phase shift Butterworth) and ground reaction forces (6 Hz, 4th order, zero-phase shift Butterworth), which became inputs to OpenSim’s Inverse Dynamics tool to determine joint moments. We found the maximum or minimum value of the first and second halves of stance phase to evaluate changes in peak sagittal plane kinematics and kinetics between baseline and feedback walking.

To analyze changes in EMG while walking with feedback, we filtered and normalized the raw EMG signals and averaged each muscle’s EMG over the stance phase. We averaged values for the medial and lateral vasti, medial and lateral gastrocnemii, and biceps femoris and semitendinosus to evaluate changes in knee-spanning muscle groups. The scalar EMG values for all steps in the baseline trial were averaged to form a baseline value for each muscle group. Then, for each muscle group and step in the feedback trials, we computed the percent change from baseline. Using a moving average with a window size of 50 steps, we identified the segment with the greatest gastrocnemius reduction. We also identified the segment that maximized gastrocnemius reductions while minimizing vasti increases by summing the gastrocnemius and vasti percent changes and selecting the 50-step window with the most negative value. Because the study aimed to determine the joint-offloading potential of the intervention, we analyzed knee contact force from steps in the segment that minimized the sum of gastrocnemius and vasti percent changes. We selected this window of steps to represent the joint loading that could be expected after a multi-day training protocol where additional learning has occurred [9].

We estimated knee contact force for five steps with gastrocnemius EMG closest to the average of each the baseline trial and the feedback trial 50-step window. We calculated muscle activations using a custom static optimization implementation. This algorithm included passive muscle forces and tendon compliance, and the objective function minimized the sum of squared muscle activations [14].

To observe the effect of reducing gastrocnemius EMG on knee contact force, it was important to capture the change in activation between the baseline and feedback static optimization solutions. Therefore, the muscle activations in the baseline trial were calculated with no EMG constraints. We added a 40 ms electromechanical delay to the processed EMG signals [27]. Then, using this baseline solution and the corresponding experimental EMG data, we calculated a scale factor for the medial and lateral gastrocnemius by dividing the static optimization activation by the measured EMG signal and averaging the factors from the five steps. We then used these factors to scale the feedback trial EMG data for the medial and lateral gastrocnemius. The static optimization function for the feedback trial was constrained to match the scaled medial and lateral gastrocnemius signals within 2%.

We then used the muscle activations and the Joint Reaction Analysis tool in OpenSim to calculate knee contact force along the longitudinal axis of the tibia. We identified early-stance and late-stance contact force peaks as the maximum value from 15– 35% and 65–85% stance, respectively, to account for variations in timing from typical 25% and 75% stance peaks. Additionally, we integrated knee contact force over stance time to find the contact force impulse.

### C. Statistics

Prior to the experiment, we calculated the sample size with a power analysis in R (v4.0.2, R Foundation for Statistical Computing, Vienna, Austria). We expected a similar effect of the intervention on knee contact force as we observed in individuals without knee osteoarthritis [14]. Using this 0.38±0.39 BW change in the late-stance peak of knee contact force, power of 0.9, and α = 0.05, we determined that 13 participants were needed for the second session.

Statistical tests were performed in MATLAB R2024a. Average muscle activity over stance phase and peaks of knee contact force, joint angles, and moments were found for each participant for baseline and feedback selected steps during the second session. We first tested for normality using the Shapiro Wilk test. If normally distributed, we compared baseline and feedback trial data using a two-sided paired t-test. If not normally distributed, we used a two-sided Wilcoxon signed rank test. Our primary outcomes were changes in gastrocnemius EMG and the late-stance peak of knee contact force. All other analyses were considered exploratory. Since the two primary outcomes were different in nature, we did not correct for multiple comparisons for these two tests. For all exploratory outcomes, we controlled for the false discovery rate using the Benjamini-Hochberg procedure [28] and we report adjusted p-values. We report values as mean ± standard deviation and α = 0.05.

## III. Results

### A. EMG

During the first session, 13 of 18 participants (72%) reduced their medial gastrocnemius EMG from baseline by an average of 10% or more. 12 of these individuals reduced their gastrocnemius EMG by 10% within the first two trials, or 12 minutes of training. In the second training session, all 13 participants again reduced their medial gastrocnemius EMG by at least 10% from baseline (Fig. 2a).

**Fig. 2.**
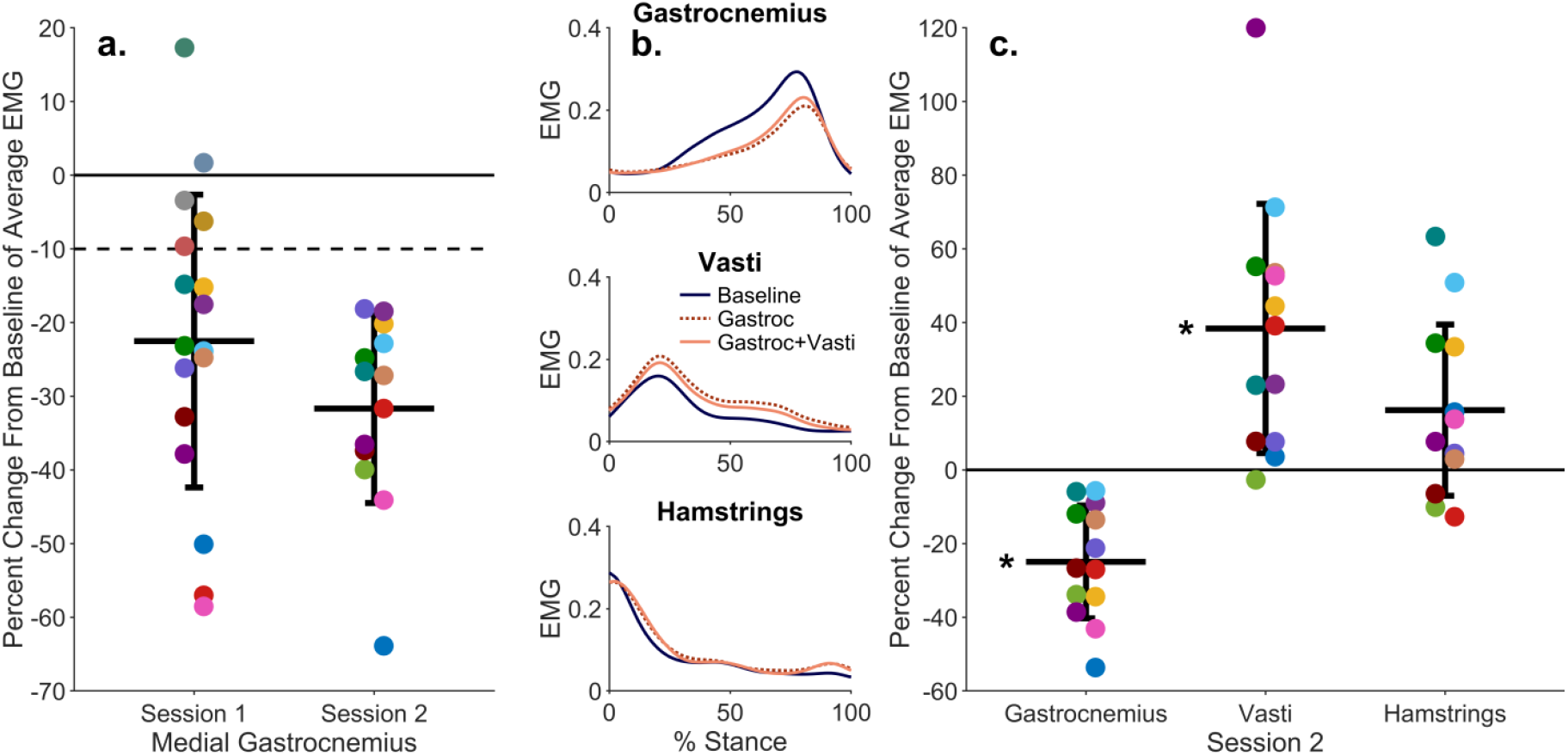
(a) Percent change from baseline of the medial gastrocnemius EMG signal during each training session, averaged over stance phase for the set of 50 consecutive steps in which the participant achieved the greatest reduction in medial gastrocnemius EMG. The horizontal bars represent the group mean, error bars represent one standard deviation, and each point is one participant (colors consistent across each plot). (b) Average EMG profiles during stance phase for the gastrocnemius, vasti, and hamstrings on the leg receiving feedback for the 13 participants in the second session. Average EMG is shown for the baseline trial, the 50 steps with the best gastrocnemius reduction (Gastroc), and the 50 steps that minimized the sum of the gastrocnemius percent change and the vasti percent change (Gastroc+Vasti). The latter set of steps was selected for further analysis. (c) Percent change from baseline of the knee-spanning muscle groups average EMG for the selected steps (Gastroc+Vasti) during the second session (*p<0.05).

During the second session, the segment of steps with the greatest gastrocnemius EMG reduction resulted in an average reduction of 28% from baseline. However, during these steps, average vasti EMG increased from baseline by 57% (Fig. 2b). The segment of steps that minimized the sum of the gastrocnemius percent change and vasti percent change resulted in a 25±15% reduction in gastrocnemius EMG from baseline (p<0.001) and a 38±34% increase in vasti EMG from baseline (p=0.004) (Fig. 2c).

After the second session, we asked participants to describe how they walked while successfully meeting the feedback goal. The most common responses were to avoid pushing off the toes and pulling the leg up from the hip. Additional strategies are included in the Supplemental Material.

We compared the measured EMG to muscle activations from static optimization. Despite magnitude differences between the EMG and activations, the activations matched the EMG waveforms for all muscles, except the rectus femoris. Similarly, changes in static optimization activations aligned with changes in EMG from the baseline to the feedback trial for the largest contributors to knee contact force: the gastrocnemius, vasti, and hamstrings (Fig. 3). The percent change from baseline to feedback trials for the gastrocnemius activation was 28±20% and 45±77% for vasti activation.

**Fig. 3.**
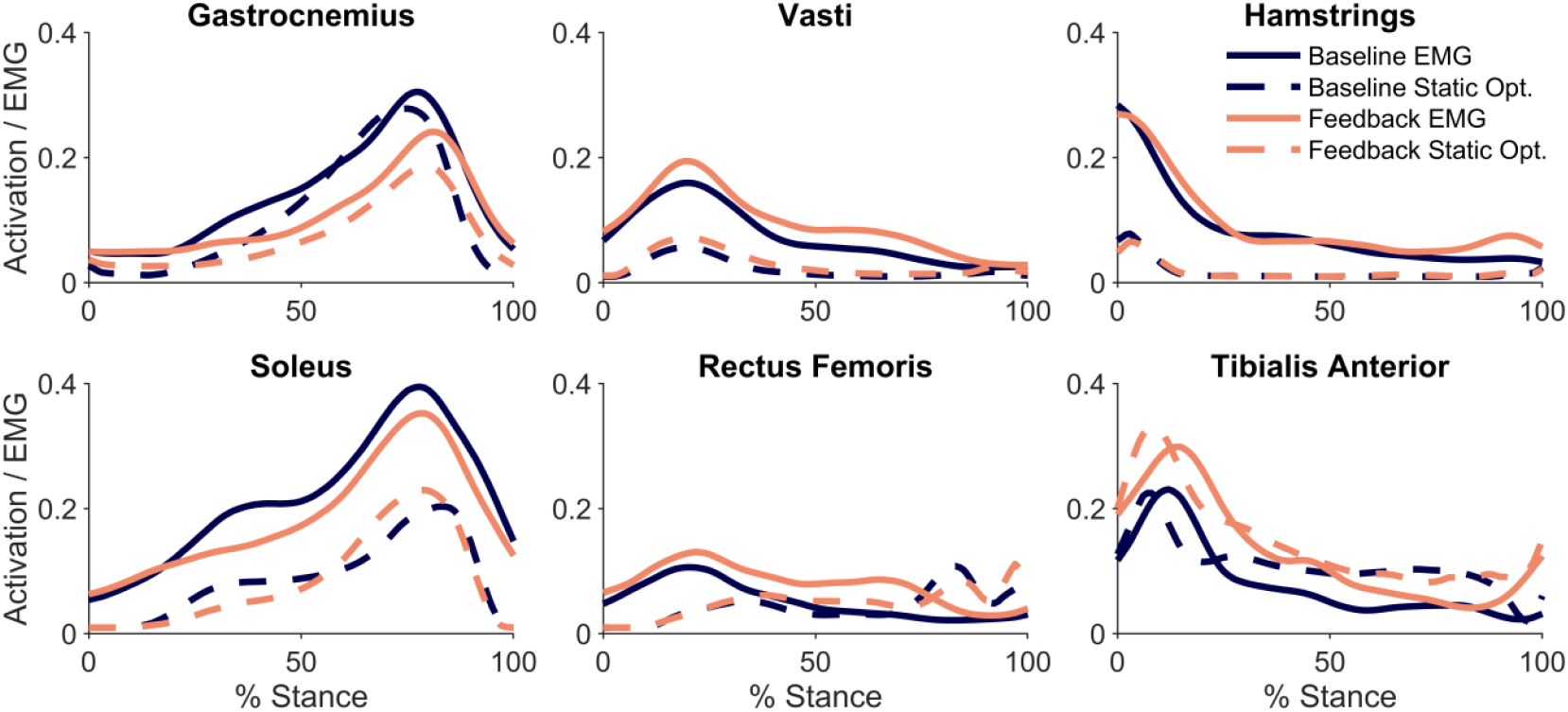
Measured EMG and muscle activation from static optimization for baseline and feedback trials, averaged across the five analyzed steps of each trial for all participants (n=13).

### B. Knee Contact Force

While walking with haptic feedback, the late-stance peak of knee contact force was reduced from baseline by an average of 0.38±0.47 BW (p=0.01), or 12% (Fig. 4). Eleven of the thirteen (85%) participants who reduced gastrocnemius EMG while walking also reduced their late-stance peak of knee contact force from baseline, but eight increased their early-stance peak. However, on average, the first peak of knee contact force did not significantly change (0.15±0.37 BW, p=0.25). Despite increases in the early-stance peak of knee contact force for some individuals, ten of thirteen (77%) reduced their contact force impulse from baseline.

**Fig. 4.**
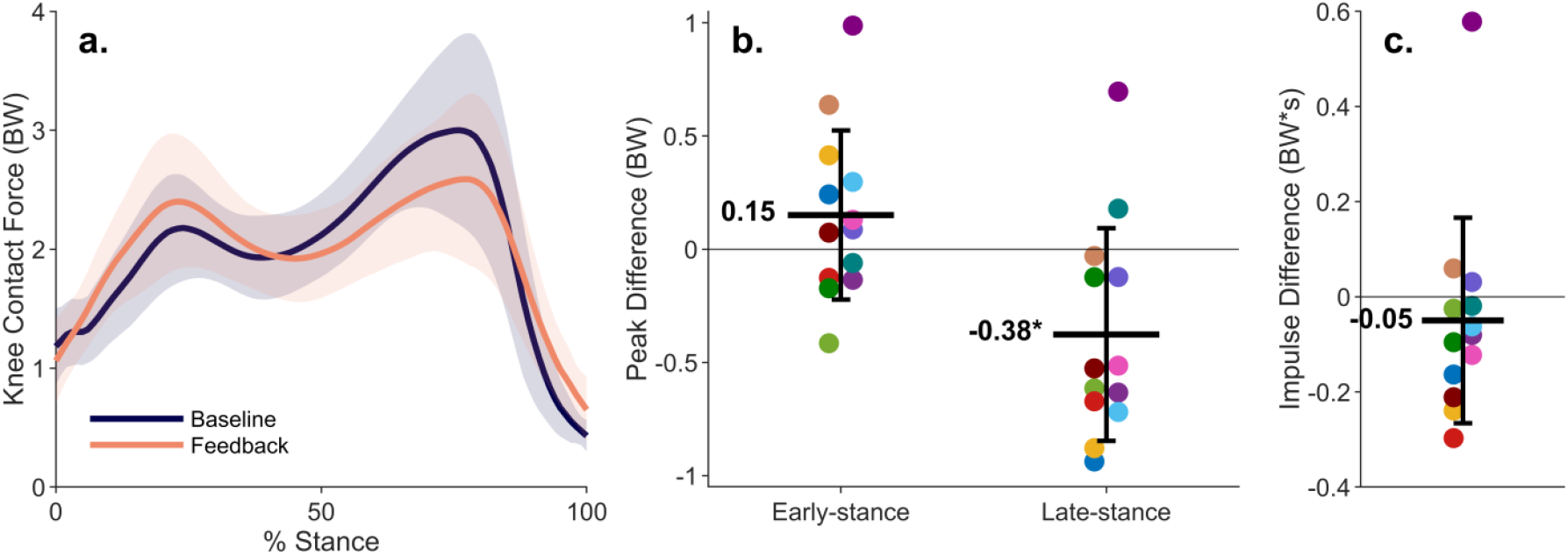
Changes in knee contact force from muscle coordination retraining. (a) Average knee contact force (line) and standard deviation (shaded region) for all participants (n=13) during baseline walking and walking with feedback. (b) Differences in knee contact force peaks for each participant between baseline and feedback walking. On average, participants reduced the late-stance peak of knee contact force from baseline by 12% (*p=0.01). (c) Difference in knee contact force impulse for each participant between baseline and feedback walking.

### C. Joint Angles and Moments

We also investigated the effect of the intervention on lower-limb sagittal plane kinematics and kinetics (Fig. 5). On average, participants walked with 3.0±3.2° (p=0.004) more ankle dorsiflexion during early stance while receiving feedback than baseline walking. The ankle plantarflexion and knee flexion moments decreased from baseline in late stance by 0.92±0.83% BW*height (p=0.007) and 0.64±0.45% BW*height (p=0.004), respectively.

**Fig. 5.**
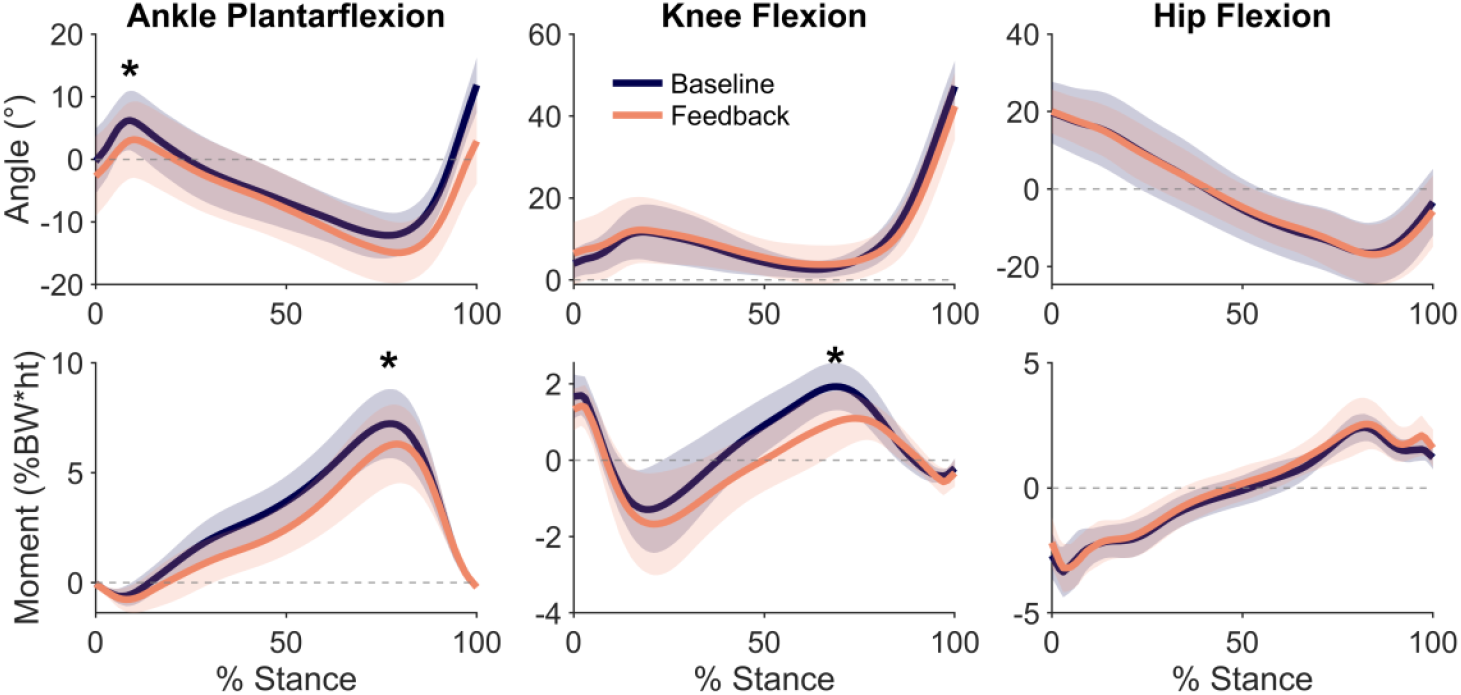
Ankle, knee, and hip angles (top row) and moments (bottom row) during baseline and feedback walking, with the mean (line) and standard deviation (shaded region) of the analyzed steps of all participants during the second session (n=13). With feedback, participants changed peak ankle dorsiflexion in early-stance and peak ankle plantarflexion and knee flexion moments in late-stance (*p<0.05).

### D. Ankle and Hip Contact Forces, Contralateral Limb

Our coordination retraining focused on reducing contact force at the knee; however, it is also important to evaluate the effects of the intervention on the hip, ankle, and contralateral limb. There were no statistically significant changes in early-stance or late-stance contact force peaks at the hip or ankle on either limb (p>0.13, Supplementary Material Fig. S1 and Fig. S3). The EMG of the contralateral medial gastrocnemius did not change from baseline, while the EMG of the contralateral vastus medialis increased by 46±29% (p=0.03) (Supplementary Material Fig. S2). This likely contributed to an increase in the first peak of contralateral knee contact force by 0.38±0.39 BW (p=0.04) (Supplementary Material Fig. S3). We observed changes in contralateral limb ankle kinematics and knee moments (Supplementary Material Fig. S4).

## IV. Discussion

The purpose of this study was to determine whether individuals with knee osteoarthritis could reduce their knee contact force while receiving haptic feedback that instructed them to reduce their gastrocnemius EMG. Thirteen of eighteen participants reduced gastrocnemius EMG in one session with feedback, and those who reduced their gastrocnemius EMG reduced their late-stance peak of knee contact force by 12%. We observed an increase in vasti muscle EMG when walking with feedback, which led to an increase in early-stance knee contact force for some individuals. Despite this, ten of thirteen individuals reduced their knee contact force impulse over the stance phase with feedback. These results demonstrate that haptic muscle coordination retraining is a promising intervention to reduce joint loads in individuals with knee osteoarthritis, though further refinement is needed to minimize the compensatory increases in vasti EMG.

Participants in this study achieved similar reductions in gastrocnemius EMG and late-stance peak knee contact force as individuals without osteoarthritis in prior work (17% and 12%, respectively) [14], despite differences in the population and feedback modality. Here, we used haptic biofeedback based only on the gastrocnemius EMG, while Uhlrich et al. [14] utilized a visual display to also incorporate feedback about the gastrocnemius to soleus EMG ratio. Achieving similar reductions in the late-stance peak of knee contact force with haptic feedback is encouraging as haptic feedback is well-suited for use in a wearable device that could deliver this coordination retraining in real-world environments [29].

Both visual and haptic feedback have previously been used in gait modifications to reduce the knee adduction moment [30] and in EMG-based gait training to improve ankle movements in children with cerebral palsy [31], [32]. Additionally, using visual feedback to increase soleus EMG has been shown to reduce the second peak of knee contact force in healthy young adults [33], providing further support for targeting muscle activity in interventions to reduce knee loading. Feedback facilitates motor learning [34], and repeated training sessions can improve retention and reduce the difficulty of the modification [7], [35]. Compared to visual feedback, haptic feedback is more effective for overground walking [36] because it can be unobtrusive and does not limit vision. Moving towards gait retraining out of the lab could expand access and utility of these interventions. Preventing compensatory increases in vasti EMG is an important direction for future work. It is possible that co-contraction contributed to the increased EMG, which can occur as people learn new gait tasks and can decrease with additional training [37]. During pilot testing, we observed that some individuals increased vasti EMG, and we attempted to reduce this effect by providing haptic feedback of both gastrocnemius and vasti EMG. However, receiving feedback about both muscles at one time made it difficult for individuals to reduce gastrocnemius EMG. Thus, for this study, we simplified the feedback to the primary goal— reducing gastrocnemius EMG. A potential solution could build on the proposed feedback by adding feedback discouraging increases in vasti EMG after individuals have learned to reduce gastrocnemius EMG, since implementing feedback on one parameter at a time has been shown to be beneficial for learning multiple tasks [38].

Musculoskeletal modeling and simulations can help discover interventions that could reduce knee contact force. Testing computationally inspired methods in experiments is necessary since human behavior does not always match modeled results. Simulations indicated that it is possible to reduce gastrocnemius activation without changing kinematics [15], [16], [17]. In this study, participants walked with a more dorsiflexed ankle during the feedback trials. Many participants reported that they avoided pushing off from their toes to meet the feedback goal, which would reduce the activity of plantarflexor muscles and aligns with our finding of a more dorsiflexed ankle. Furthermore, reductions in late-stance ankle and knee moments suggest that redundant muscles, the soleus and hamstrings, did not fully compensate for the reduced gastrocnemius force. This differs from our previous study where we provided feedback to reduce gastrocnemius EMG and increase soleus EMG, which reduced knee contact force without reducing the ankle plantarflexion moment [14]. Here, we prioritized making the feedback as simple as possible to promote reductions in gastrocnemius EMG and knee contact force. Both feedback approaches resulted in similar reductions in late-stance knee flexion moments and late-stance knee contact force.

We provided unilateral biofeedback and instructed participants to walk symmetrically. However, despite similar changes in ankle kinematics on both limbs, participants did not reduce contralateral gastrocnemius EMG (Supplementary Material Fig. S2). Participants increased contralateral vasti EMG, which likely contributed to an increase in the early-stance peak of knee loading on the contralateral leg (Supplementary Material Fig. S3). These findings indicate that bilateral feedback may be necessary, especially for individuals with bilateral knee osteoarthritis.

Most of our participants were able to reduce their gastrocnemius EMG by at least 10% within the first training session, but some (28%) were unable to reach this threshold within 30 minutes of training. More individuals may be able to alter their gastrocnemius coordination with additional training; however, our results show that 92% of those who could reduce their gastrocnemius EMG did so within the first 12 minutes. Thus, a rapid initial screening session may be practical to determine whether an individual can reduce their gastrocnemius EMG and reduce knee contact force prior to prescribing the intervention. Similarly, understanding the effects of the coordination retraining on other lower-limb joints can help ensure the intervention is beneficial for those who adopt it. On average, we found no change from baseline in hip or ankle contact force on either limb (Supplementary Material Fig. S1and Fig. S3). However, these contact forces increased for some participants. On the other hand, some participants reduced hip contact force; thus, muscle coordination strategies could be developed to reduce both knee and hip loading [16]. Given the participant-specific effects of the intervention in ipsilateral and contralateral joint loading, it will likely be important to evaluate how each participant’s joint loading responds to the coordination retraining before suggesting long-term adoption of the modification to avoid unintended increases in loading [39]. A biomechanical screening procedure is becoming more clinically feasible with advances in mobile sensing technologies for rapidly estimating joint loading (e.g., wearable sensors and videos) [40], [41].

It is important to acknowledge the limitations of our study. First, knee contact forces can only be directly measured using an instrumented knee implant, so musculoskeletal models are necessary to estimate internal joint forces. However, the static optimization method we used has been shown to accurately detect reductions in late-stance knee contact force induced by gait retraining interventions in instrumented knees as small as 0.05 times body weight [42], which is smaller than our average 0.38 body weight contact force reduction. Second, we analyzed sections of trials that maximized the reduction in gastrocnemius EMG while minimizing the increase in vasti EMG. This selection may produce more favorable force-reduction estimates, and a different selection would likely result in a larger increase in early-stance knee contact force than we report. However, we were interested in identifying the changes in knee contact force that may be possible after longer term training where we would expect inefficient increases in vasti EMG to subside [37]. Additionally, we considered average modifications maintained through sections of trials by averaging over 50-step windows. It is reasonable to assume that individuals would be able to achieve at least the reported levels of contact force changes with additional training and vasti EMG feedback. This study did not evaluate retention of the coordination changes without feedback, limiting its generalizability beyond the short in-lab training sessions. Future studies are needed to determine the amount of training necessary to maintain reductions in gastrocnemius EMG and late-stance knee contact force. Our study included individuals with a wide range of osteoarthritis severity, KL grades 1–4, who were all able to reduce their gastrocnemius EMG while walking. We did not evaluate KL grade for those who did not reduce gastrocnemius EMG, but future studies may consider investigating response to the intervention based on severity, since those with more severe radiographic osteoarthritis may be less responsive to offloading interventions [43]. Additionally, we did not evaluate changes in pain and function, but future studies should focus on these clinical outcomes. The reductions in knee contact force from this study warrant longitudinal clinical studies, as their magnitude is comparable to that achieved through weight loss interventions shown to reduce pain and improve function [44], [45], [46].

## V. Conclusion

We found that walking with haptic biofeedback to reduce gastrocnemius EMG can reduce the late-stance peak of knee contact force for individuals with knee osteoarthritis. Although we observed an increase in vasti EMG and the early-stance peak of knee contact force in some individuals, most participants reduced their knee contact force impulse. While additional studies are needed, this work demonstrates the promise of low-cost, non-surgical interventions—such as muscle coordination retraining—to address a critical treatment gap for individuals with mild-to-moderate knee osteoarthritis.

## Data Availability

All data produced in the present study are available upon reasonable request to the authors.

## Acknowledgment

S.L.D and S.D.U are inventors on a granted U.S. patent (US-12193808-B2) related to the muscle coordination retraining method, entitled “Real-time electromyography feedback to change muscle activity during complex movements.” The other authors declare no competing interests. We thank Josué Gil-Silva for help in processing motion capture data.

