## Supplementary Material for "Retraining Gastrocnemius Muscle Coordination Reduces Late-Stance Knee Contact Force in Individuals with Knee Osteoarthritis"

### I. Explanation of Strategy Suggestions

Before beginning the coordination retraining, we explained the function of the gastrocnemius muscle, the rationale for decreasing its activity and provided strategy suggestions to help participants reduce their gastrocnemius electromyography (EMG). These suggestions arose from effective strategies identified during pilot testing. We explained the first two suggestions to all participants at the start, and if another suggestion was needed after a feedback trial, we read the third.

1. "Focus on using your hip muscles to move your leg through each step, instead of your calf muscles. When you are about to take a step off the ground, try to feel your leg being pulled up from your hip instead of pushed up from your foot."
2. "Keep more of your weight in the heel of your foot while walking, and avoid going up onto your toes for a large push off the ground. Try to have less force in the front of your foot while walking."
3. "Try to have more of your stride in front of you, and lead with your legs, rather than your upper body. There are a few options to try to do this. You can reach your heels further forward, lean slightly back, or tuck your tailbone. You can try any of these or combine them, whatever is most comfortable and effective for you. Keep your legs and ankles relaxed."

### II. Strategies Used

After completing the second session, we asked participants to respond to the following prompt: "Describe how you walked when you were able to meet the goal to stop the feedback." Table S1 lists the most effective strategies participants reported and the number of participants who included the strategy in their response. Many participants used combinations of the strategies, so the count of responses is greater than number of participants.

TABLE SI
BEST STRATEGIES USED BY PARTICIPANTS DURING SESSION 2

| Strategy | Responses |
|---|---|
| Avoid pushing off from toes (lifting toes up, keeping weight in heels) | 8 |
| Pull leg up from the hip | 5 |
| Engage core muscles | 4 |
| Lean slightly back, shift center of mass back | 3 |
| Stand up straighter | 3 |
| Tilt pelvis back | 2 |

### III. Supplemental Results

Additional exploratory analyses about hip and ankle contact forces, the contralateral limb, and ground reaction forces are included here. For all comparisons between the baseline and feedback trials, we used a Shapiro-Wilk test to first check for normality, and according to the result, compared changes using a two-sided paired t-test or a two-sided Wilcoxon signed rank test. We controlled for the false discovery rate with the Benjamini-Hochberg procedure for the supplementary analyses as a group (37 tests). We report adjusted p-values, $\alpha=0.05$, and values as mean ± standard deviation.

#### *A. Hip and Ankle Contact Force*

We evaluated changes in joint contact force at the hip and ankle from the gastrocnemius coordination retraining. Hip contact force was calculated as the force along the long axis of the femur. Ankle contact force was calculated as the force along the inferior-superior axis of the talus. Using the same analyzed steps as the knee contact force results, we similarly found peaks for each participant ($n=13$) between 15 and 35% of stance and 65 to 85% of the stance phase for the baseline and feedback trials.

Across the group, there were no significant changes in the early or late stance peaks of hip or ankle contact force between conditions ($p>0.17$) (Fig. S1). However, some participants did increase contact force at the hip or ankle.

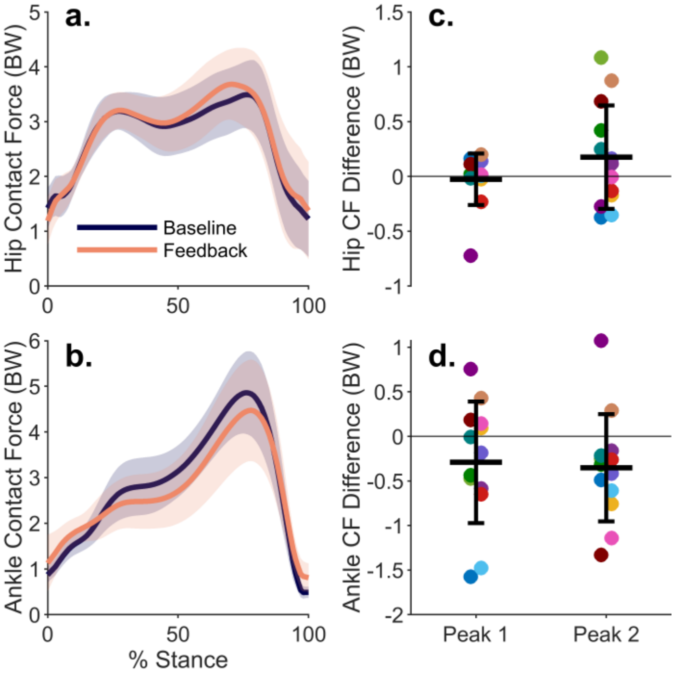


Fig. S1. Average (a) hip and (b) ankle contact force (line) and standard deviation (shaded region) for all participants (n=13) during baseline walking and walking with feedback. Differences in (c) hip and (d) ankle contact force (CF) peaks for each participant between baseline walking and walking with feedback. There were no statistically significant changes in any of these contact force peaks (p>0.16).

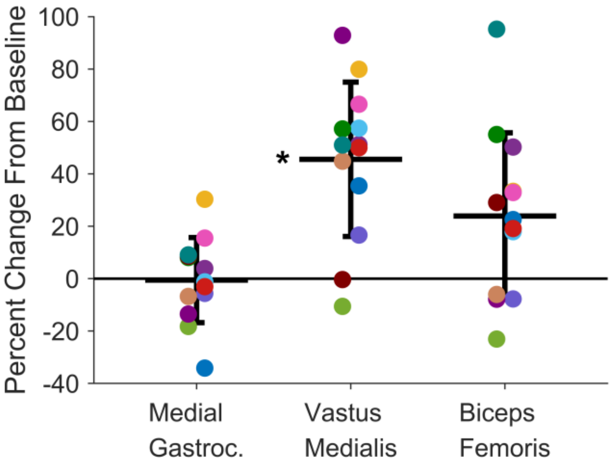


Fig. S2. Percent change from baseline of the average stance-phase EMG of the knee-spanning muscles measured on the contralateral limb, from the window of 50 steps corresponding to the steps analyzed on the leg receiving feedback during the second session. The horizontal bars represent the group mean, error bars represent ± one standard deviation, and each point is one participant (n=13).

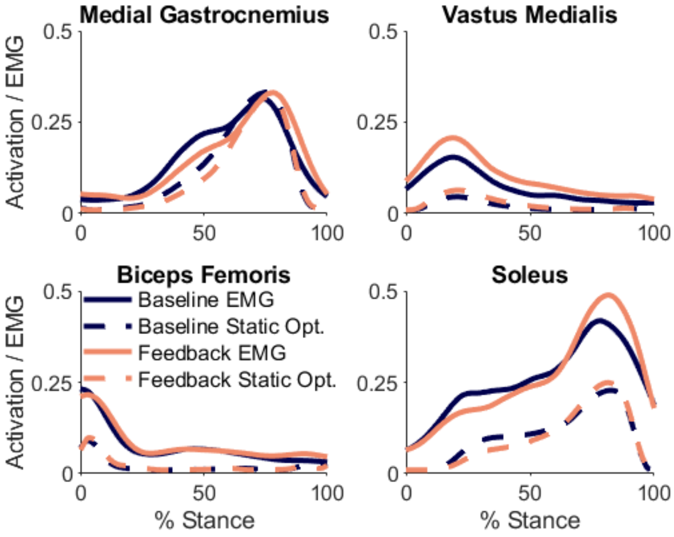


Fig. S3. Measured EMG and muscle activation from static optimization on the contralateral limb for baseline and feedback trials, averaged across the five analyzed steps of each trial for all participants (n=13).

### *B. Effects on the contralateral limb*

During the feedback trials, participants received haptic biofeedback on one leg. We instructed participants to walk symmetrically; however, it is important to evaluate changes on the leg that did not receive feedback (i.e., contralateral limb). We evaluated EMG, joint contact forces, kinematics, and kinetics.

#### *1) EMG*

We analyzed EMG data for the contralateral limb from the medial gastrocnemius, vastus medialis, and biceps femoris. We calculated the percent change from baseline for the same window of 50 steps analyzed on the feedback limb during the second session.

On average, vasti EMG increased by 46±29% (p=0.03) from baseline (Fig. S2).

#### *2) Knee, Hip, and Ankle Contact Force*

To evaluate changes in knee, hip, and ankle contact force on the contralateral limb, we analyzed five steps from the baseline trial and five steps from the feedback trial corresponding to the steps analyzed on the ipsilateral limb. We utilized the same static optimization implementation as described in the Methods section, except no EMG constraints were applied in either baseline or feedback simulations of the contralateral limb. EMG constraints were not used because the contralateral leg did not receive feedback and only had EMG on the medial, not lateral, gastrocnemius. Despite this, the muscle activation waveforms and relative changes between conditions are similar to the measured EMGs (Fig. S3). There is a 40 ms electromechanical delay added to the EMG signals. Contact force peaks were identified at each joint for each participant in early-stance (15-35% stance) and late-stance (65-85% stance) and compared between baseline and feedback trials.

The first peak of contralateral knee contact force increased by 0.38±0.39 times body weight (p=0.04) (Fig. S4), which was likely influenced by the observed increase in vasti EMG. There were no significant changes in the second peak of knee contact force or either peak of hip or ankle contact force (p>0.13). Despite this, it is important to consider individual changes—one participant experienced a 0.6 body weight increase in late-stance contact force in the contralateral knee.

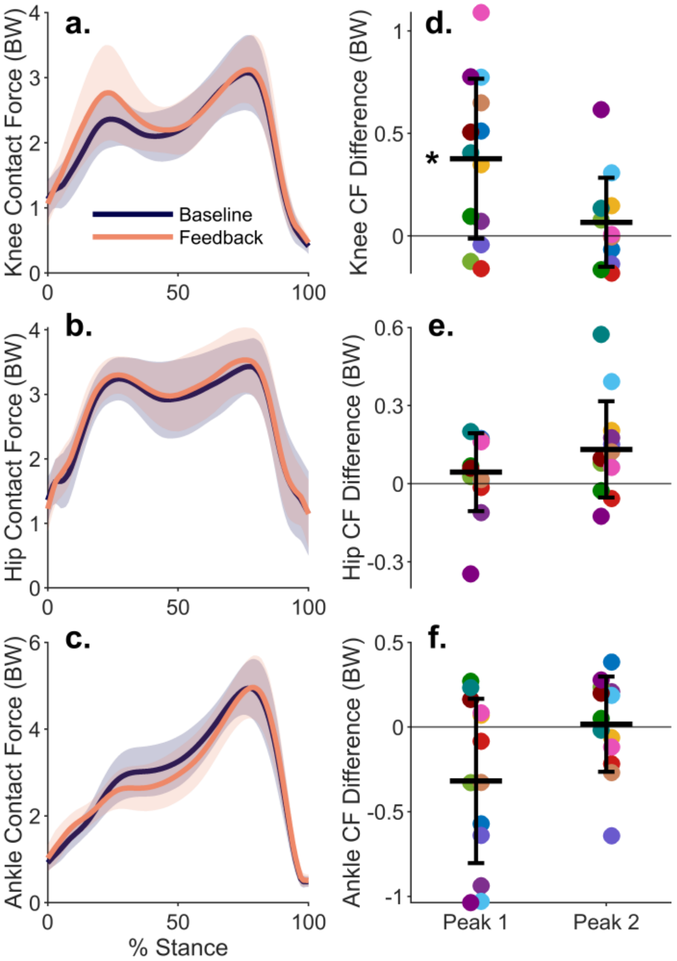


Fig. S4. Average contralateral (a) knee, (b) hip, and (c) ankle contact force (line) and standard deviation (shaded region) for all participants (n=13) during baseline walking and walking with feedback. Differences in (d) knee, (e) hip, and (f) ankle contact force (CF) peaks for each participant between baseline walking and walking with feedback. (*p<0.05)

#### 3) *Kinematics and Kinetics*

We additionally evaluated changes in kinematics and kinetics on the contralateral limb during the second session of coordination retraining. We averaged joint angles and moments for the analyzed steps for each participant (n=13) and identified the maximum or minimum value of the first and second halves of the stance phase to compare peaks between baseline and feedback trials.

Ankle kinematics changed similarly on the contralateral leg to the ipsilateral limb, with 3.5±3.1° more dorsiflexion in early-stance (p=0.03, Fig. S5). The late-stance knee flexion moment decreased by 0.48±0.47% BW*height (p=0.01).

### C. *Ground Reaction Forces*

We compared ground reaction forces for the ipsilateral and contralateral legs (Fig. S6) and found no changes (p>0.12) at the peaks between baseline and feedback trials during the analyzed steps.

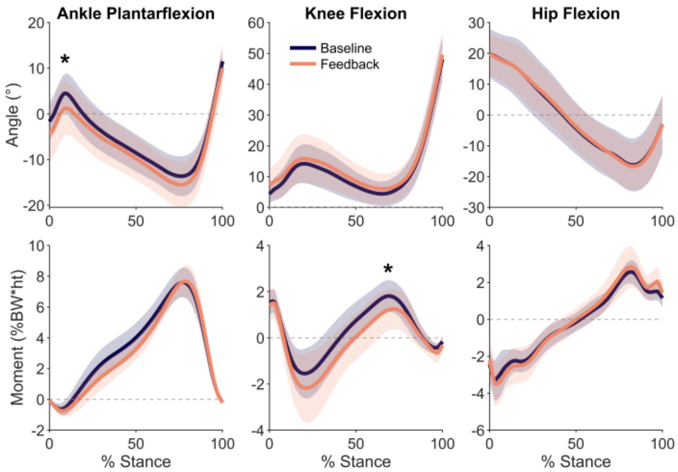


Fig. S5. Contralateral limb ankle, knee, and hip angles (top row) and moments (bottom row) during baseline and feedback walking, with the mean (line) and standard deviation (shaded region) of the analyzed steps of all participants during the second session (n=13). (*p<0.05)

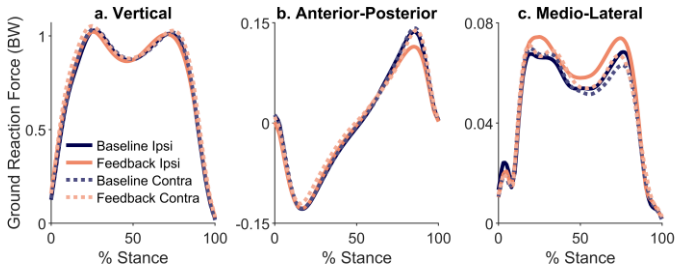


Fig. S6. Mean (a) vertical, (b) anterior-posterior, and (c) medio-lateral ground reaction forces for both limbs during baseline and feedback walking of the analyzed steps of all participants during the second session (n=13).

### IV. Average EMG for All Measured Muscles

We recorded EMG from 13 muscles during the second session, nine on the leg receiving feedback and four on the contralateral limb. The filtered and normalized signals during stance phase were averaged for the 13 participants for the five analyzed steps of each the baseline and feedback trial (Fig. S7).

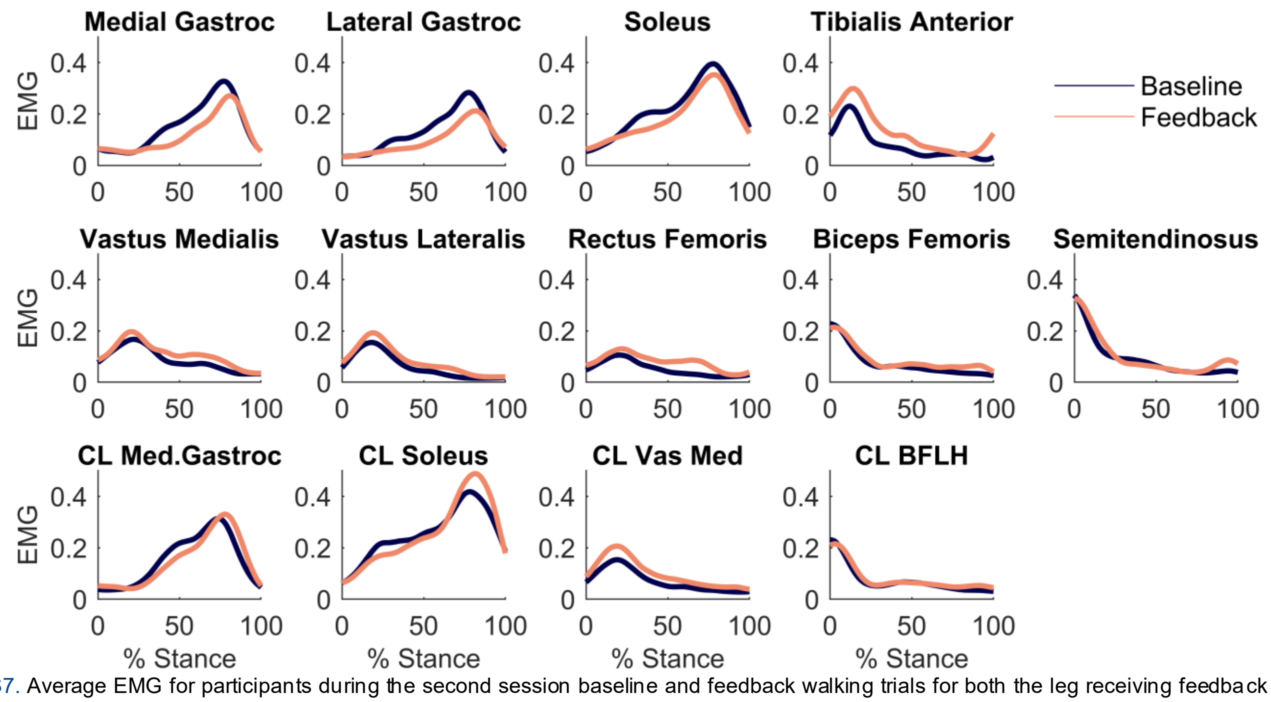


Fig. S7. Average EMG for participants during the second session baseline and feedback walking trials for both the leg receiving feedback and a subset of muscles on the contralateral limb (CL).

### V. Tables of Results

TABLE S2
RESULTS FOR ALL METRICS COMPUTED FOR THE IPSILATERAL LIMB

| IPSILATERAL LIMB | MEAN DIFFERENCE | STANDARD DEVIATION | P-VALUE | EFFECT SIZE |
|---|---|---|---|---|
| ***EMG*** | *Normalized EMG (% Change)* | | | |
| Gastrocnemius | -0.07 (-25) | 0.04 (15) | <0.001 | -1.60 |
| Vasti | 0.04 (38) | 0.03 (34) | 0.004 | 1.24 |
| Hamstring | 0.02 (16) | 0.04 (23) | 0.12 | 0.58 |
| ***Knee Contact Force*** | *BW* | | | |
| Early-stance Peak | 0.15 | 0.37 | 0.25 | 0.40 |
| Late-stance Peak | -0.38 | 0.47 | 0.01 | -0.8 |
| Impulse | -0.05 *BW*s* | 0.22 | 0.14 | -0.23 |
| ***Joint Angles*** | *Degrees* | | | |
| Early-stance Ankle Plantarflexion | -3.04 | 3.22 | 0.004 | -0.94 |
| Late-stance Ankle Plantarflexion | -2.79 | 3.88 | 0.07 | -0.72 |
| Early-stance Knee Flexion | 0.41 | 5.13 | 0.25 | 0.08 |
| Late-stance Knee Flexion | 1.15 | 4.66 | 0.84 | 0.25 |
| Early-stance Hip Flexion | 0.26 | 5.16 | 0.40 | 0.05 |
| Late-stance Hip Flexion | -0.48 | 5.86 | 0.34 | -0.08 |
| ***Joint Moments*** | *% BW*height* | | | |
| Early-stance Ankle Plantarflexion | -0.14 | 0.25 | 0.12 | -0.57 |
| Late-stance Ankle Plantarflexion | -0.92 | 0.83 | 0.007 | -1.11 |
| Early-stance Knee Flexion | -0.41 | 1.06 | 0.25 | -0.39 |
| Late-stance Knee Flexion | -0.64 | 0.45 | 0.004 | -1.42 |
| Early-stance Hip Flexion | 0.16 | 0.52 | 0.34 | 0.30 |
| Late-stance Hip Flexion | 0.28 | 0.58 | 0.07 | 0.49 |
| ***Hip Contact Force*** | *BW* | | | |

| | | | | |
|---|---|---|---|---|
| Early-stance Peak | -0.03 | 0.24 | 0.79 | -0.11 |
| Late-stance Peak | 0.18 | 0.47 | 0.34 | 0.37 |
| ***Ankle Contact Force*** | *BW* | | | |
| Early-stance Peak | -0.29 | 0.68 | 0.29 | -0.43 |
| Late-stance Peak | -0.35 | 0.60 | 0.17 | -0.58 |
| ***Ground Reaction Forces*** | *BW* | | | |
| Early-stance Vertical | -0.02 | 0.03 | 0.21 | -0.53 |
| Late-stance Vertical | -0.01 | 0.03 | 0.29 | -0.43 |
| Early-stance Anterior-Posterior | 0.002 | 0.02 | 0.84 | 0.07 |
| Late-stance Anterior-Posterior | -0.02 | 0.03 | 0.14 | -0.64 |
| Early-stance Medio-Lateral | 0.004 | 0.02 | 0.59 | 0.21 |
| Late-stance Medio-Lateral | 0.006 | 0.01 | 0.14 | 0.67 |

TABLE S3

RESULTS FOR ALL METRICS COMPUTED FOR THE CONTRALATERAL LIMB

| CONTRALATERAL LIMB | MEAN DIFFERENCE | STANDARD DEVIATION | P-VALUE | EFFECT SIZE |
|---|---|---|---|---|
| ***EMG*** | *Normalized EMG (% Change)* | | | |
| Gastrocnemius | -0.01 (-0.56) | 0.02 (16) | 0.84 | -0.28 |
| Vasti | 0.03 (46) | 0.03 (29) | 0.03 | 0.91 |
| Hamstring | 0.01 (24) | 0.03 (32) | 0.23 | 0.19 |
| ***Knee Contact Force*** | *BW* | | | |
| Early-stance Peak | 0.38 | 0.39 | 0.04 | 0.97 |
| Late-stance Peak | 0.07 | 0.22 | 0.43 | 0.31 |
| ***Joint Angles*** | *Degrees* | | | |
| Early-stance Ankle Plantarflexion | -3.48 | 3.10 | 0.03 | -1.12 |
| Late-stance Ankle Plantarflexion | -1.80 | 2.88 | 0.15 | -0.62 |
| Early-stance Knee Flexion | 1.62 | 2.97 | 0.21 | 0.55 |
| Late-stance Knee Flexion | 1.58 | 3.60 | 0.42 | 0.44 |
| Early-stance Hip Flexion | 0.01 | 4.41 | 0.43 | 0.003 |
| Late-stance Hip Flexion | -0.57 | 4.11 | 0.29 | -0.14 |
| ***Joint Moments*** | *% BW*height* | | | |
| Early-stance Ankle Plantarflexion | -0.08 | 0.22 | 0.34 | -0.37 |
| Late-stance Ankle Plantarflexion | 0.07 | 0.44 | 0.67 | 0.16 |
| Early-stance Knee Flexion | -0.64 | 0.72 | 0.06 | -0.89 |
| Late-stance Knee Flexion | -0.48 | 0.47 | 0.01 | -1.02 |
| Early-stance Hip Flexion | -0.13 | 0.59 | 0.59 | -0.21 |
| Late-stance Hip Flexion | 0.29 | 0.71 | 0.30 | 0.41 |
| ***Hip Contact Force*** | *BW* | | | |
| Early-stance Peak | 0.04 | 0.15 | 0.25 | 0.30 |
| Late-stance Peak | 0.13 | 0.18 | 0.13 | 0.71 |
| ***Ankle Contact Force*** | *BW* | | | |
| Early-stance Peak | -0.32 | 0.49 | 0.14 | -0.66 |
| Late-stance Peak | 0.02 | 0.28 | 0.86 | 0.06 |
| ***Ground Reaction Forces*** | *BW* | | | |
| Early-stance Vertical | 0 | 0.05 | 0.98 | 0.01 |
| Late-stance Vertical | 0.02 | 0.02 | 0.12 | 0.75 |
| Early-stance Anterior-Posterior | 0.007 | 0.02 | 0.34 | 0.37 |
| Late-stance Anterior-Posterior | -0.003 | 0.01 | 0.59 | -0.20 |
| Early-stance Medio-Lateral | 0.001 | 0.01 | 0.84 | 0.09 |
| Late-stance Medio-Lateral | 0.003 | 0.01 | 0.59 | 0.22 |
